# Accuracy of rapid antigen testing for COVID-19 in shelter settings

**DOI:** 10.1101/2025.01.23.25320556

**Authors:** Yasmin Garad, Andreea A. Manea, Negin Pak, Bronwyn Barker, Danielle Kasperavicius, Lames Danok, Stefan Baral, Aaron M. Orkin, Amna Siddiqui, Sharon E. Straus, Christine Fahim

## Abstract

**Background:** The COVID-19 pandemic disproportionally affected congregate living settings, including shelters. COVID-19 transmission can have more adverse outcomes in these settings due to the vulnerability of residents. Point of care rapid antigen testing (RAT) represents a strategy with potential benefits for COVID-19 detection in shelters, including rapid results, ease of use, cost-effectiveness, and early detection.

**Objectives:** Our primary objective was to assess the real-world test accuracy of RAT for COVID-19 using the Quidel Sofia 2 Flu + SARS Antigen fluorescent immunoassay (Sofia RAT) compared to polymerase chain reaction (PCR) testing among shelter residents in Ontario, Canada.

**Study Design:** A consecutive sample of 102 residents across six shelters who were symptomatic for, or exposed to COVID-19 were included. The RAT and PCR samples were taken on the same day for each participant. Results from the Sofia RAT were compared to PCR test results to determine test accuracy. Participant demographic data could not be collected due to workforce constraints.

**Results:** We reported our methods and findings using the QUality Assessment tool of Diagnostic Accuracy Studies (QUADAS-2) guidelines. Sofia 2 RAT specificity was 97.9% (95% CI: 92.7% to 99.7%) for COVID-19 compared to PCR. Due to a lack of true positive cases, sensitivity could not accurately be calculated (0.00% (95% CI: 0.00% to 52.2%)).

**Conclusion:** These data suggest that the Sofia RAT is a highly specific test for COVID-19.

## Background

COVID-19 disproportionally affected individuals in homeless shelter settings^1,2^. Individuals in these settings experience a greater risk of physical frailty, comorbidity, and being immunocompromised than the general population^2-5^. These risk factors in combination with crowded conditions likely contributed to higher rates of COVID-19 in these settings^2,6^.

Point of care rapid antigen testing (RAT) provides multiple benefits for COVID-19 detection compared to laboratory-based testing such as polymerase chain reaction (PCR)^7^. It is a streamlined alternative, which compared to laboratory-based testing typically has shorter processing times, does not require specific expertise for sample collection and handling, and is more cost-effective^7-10^.

During the COVID-19 pandemic, RAT was used as a low-barrier and cost-effective tool for early detection^11,12^. As the pandemic evolved, new multiplex RAT point of care testing became available including the Sofia 2 Flu + SARS Antigen fluorescent immunoassay (FIA; hereafter Sofia RAT) developed by Quidel Corporation^13^. Due to its ability to simultaneously detect respiratory infections that share similar clinical presentations, such as COVID-19 and influenza, multiplex RAT can help to facilitate a streamlined approach to surveillance and diagnosis in shelter settings^14^. To our knowledge, there are limited data regarding the performance of Sofia RAT compared to PCR testing for COVID-19 in shelter settings.

### Objectives

Our main objective was to assess the real-world test accuracy of RAT for COVID-19 using the Sofia RAT compared to PCR testing among individuals in homeless shelters.

### Study Design

We reported our methods and findings in accordance with the QUality Assessment tool of Diagnostic Accuracy Studies (QUADAS-2)^15^. The project was conducted through St. Michael’s Hospital (Toronto, Ontario) from October 2022 until March 2023. Ethical approval was obtained from the Unity Health Toronto Research Ethics Board (REB# 21-319).

A consecutive sample of residents across six shelters in Toronto, Ontario who were symptomatic or exposed to COVID-19 were included. Shelters offer temporary accommodation and support services to people experiencing homelessness (PEH). Participants were recruited by nursing staff who conducted routine rapid antigen testing across the six shelters. Residents who expressed interest and provided verbal consent were included in the study and compensated with a $20 (CAD) gift card for their participation.

Multiplex RAT was implemented, which used immunofluorescence technology to simultaneously detect nucleocapsid protein from influenza A, influenza B and SARS-CoV-2. The Sofia RAT was used in combination with the benchtop Sofia 2 FIA analyzer^13,16,17^. This analyzer generated results in 15 minutes^17^.

Following informed verbal consent from participants, PCR samples were collected through a nasal or nasopharyngeal swab by a registered nurse at the same encounter where the Sofia RAT was conducted. The PCR sample was transported to St. Michael’s Hospital for analysis within 48 hours of sample collection. The results were then reported to the study team by the St. Michael’s Hospital microbiology lab by fax and were also publicly reported to Toronto Public Health, in accordance with provincial reporting requirements^18^. Positive results were reported to the participant within 72 hours post-collection.

Statistical analyses were conducted using Stata version 18^19^. All measures of diagnostic accuracy were estimated using Stata’s diagti command.

### Results

In total, 102 individuals participated in the study. Sofia 2 RAT specificity was found to be 97.9% (95% CI: 92.7% to 99.7%) (see Table 1). The percent of negative test results that were correctly identified (NPV) was 95.0% (88.7% to 98.4%) (see Table 1). Due to a lack of positive cases, sensitivity and positive predictive value could not be accurately calculated.

**Table 1.**
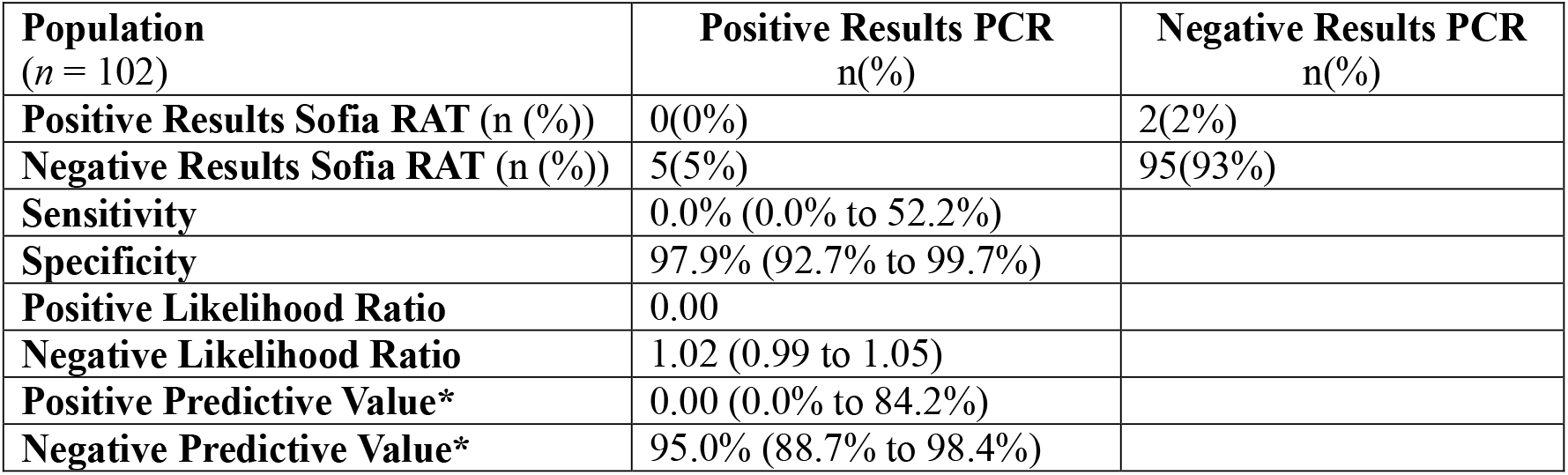
Comparison of Sofia RAT performance with PCR.

| Population<br>(*n* = 102) | Positive Results PCR<br>n(%) | Negative Results PCR<br>n(%) |
| --- | --- | --- |
| Positive Results Sofia RAT (n (%)) | 0(0%) | 2(2%) |
| Negative Results Sofia RAT (n (%)) | 5(5%) | 95(93%) |
| Sensitivity | 0.0% (0.0% to 52.2%) |  |
| Specificity | 97.9% (92.7% to 99.7%) |  |
| Positive Likelihood Ratio | 0.00 |  |
| Negative Likelihood Ratio | 1.02 (0.99 to 1.05) |  |
| Positive Predictive Value* | 0.00 (0.0% to 84.2%) |  |
| Negative Predictive Value* | 95.0% (88.7% to 98.4%) |  |

## Discussion

We determined that among individuals who were exposed or symptomatic for COVID-19, real world specificity for case detection using the Sofia RAT compared to PCR was high^20^. These results on specificity for the Sofia RAT meet the criteria set by the World Health Organization (WHO) to be considered an acceptable substitute to PCR testing (i.e. sensitivity ≥ 80% and specificity ≥ 97%)^21^ in real world settings. However, we were unable to determine reliable results for sensitivity due to a lack of positive test results in our sample. Due to this, the conclusions that can be drawn regarding the application of the Sofia RAT based on the findings of this study are limited.

Our findings on specificity for the Sofia RAT are also aligned with those of a recent Cochrane review which demonstrated that the Sofia RAT was one of only two RAT tests that met the WHO standards for specificity regardless of whether participants were symptomatic (i.e., 99.4% (98.7% to 99.8%)) or asymptomatic (i.e., 99.7% (99.3% to 99.8%))^22-28^. Several other studies similar to ours have demonstrated high specificity over 95% for COVID-19 for RAT using the Sofia 2 analyzer^22-29^. However, to our knowledge no previous studies have assessed the accuracy of the Sofia RAT in shelter settings.

In the Cochrane review noted above, the Sofia RAT was also one of seven rapid antigen tests that met acceptable performance standards for sensitivity for symptomatic participants (i.e., 80.0% (71.5% to 86.4%)) but did not meet these standards for asymptomatic individuals (i.e., 41.2% (18.4% to 67.1%). Rates of sensitivity for COVID-19 using the Sofia-2 RAT vary widely in the literature^22-28^. Given the lack of routine and rapid access to PCR testing in shelter settings and the option to use the Sofia-2 Analyzer to test for multiple infectious diseases, this test may be a useful option to support the early detection of infectious disease in these settings^30,31^.

Given its high specificity, this diagnostic tool is unlikely to have many false positives meaning that there is a low risk of diagnostic delay^32^. Furthermore, this decreased diagnostic delay can improve outbreak response, including individual clinical response like linkage to treatment, but also public health response including supportive isolation. Overall, despite the risk of false negatives, the use of rapid testing as a first step may be effective in reducing costs, laboratory staff burden, and the risk of transmission by detecting some cases quickly and implementing infection prevention and control practices^7,9,22^.

This study has some limitations. First, due to a lack of positive cases we were unable to determine a reliable estimate of sensitivity. Second, due to a lack of feasibility we were not able to distinguish between symptomatic and asymptomatic participants or measure the viral load for each sample, which may have impacted test accuracy^33^. Lastly, for similar reasons, we were not able to collect demographic data. However, our findings reflect the real-world circumstances under which diagnostic testing occurs. Despite these limitations, our findings still show high levels of accuracy in shelter settings for detecting the absence of COVID-19 using the Sofia RAT.

## Conclusion

Findings from our study demonstrated high specificity for COVID-19 testing using the Sofia RAT, however we were unable to reliably report sensitivity due to the lack of true positive cases. Individuals in shelter settings may be at greater risk of infection due to age, comorbid conditions, and/or precarious housing^3^. Multiplex testing using rapid tests such as the Sofia RAT in homeless shelter settings can support the rapid differential diagnosis of infectious diseases with similar symptoms, allowing for appropriate infection prevention and control measures to be taken more quickly^14^.

## Data Availability

The datasets generated for this study are not publicly available to protect the privacy of participants and related REB confidentiality requirements. De-identified can be made upon request.

## Acknowledgements

The authors would like to express their gratitude to all subjects for their engagement in this study. We would also to thank Elizabeth Birkurovitz, Tina Kaur, Princilla Agyemang, Grace Richandi for their contributions to the data collection for this study and manuscript review. SES was funded by a Tier 1 Canada Research Chair.

